# Serological responses to SARS-CoV-2 following non-hospitalised infection: clinical and ethnodemographic features associated with the magnitude of the antibody response

**DOI:** 10.1101/2020.11.12.20230763

**Authors:** Adrian M Shields, Sian E Faustini, Marisol Perez-Toledo, Sian Jossi, Joel D Allen, Saly Al-Taei, Claire Backhouse, Lynsey Dunbar, Daniel Ebanks, Beena Emmanuel, Aduragbemi A Faniyi, Mark I. Garvey, Annabel Grinbergs, Golaleh McGinnell, Joanne O’Neill, Yasunori Watanabe, Max Crispin, David. C Wraith, Adam F Cunningham, Mark T Drayson, Alex G Richter

## Abstract

**Objective:** To determine clinical and ethnodemographic correlates of serological responses against the SARS-CoV-2 spike glycoprotein following mild-to-moderate COVID-19.

**Design:** A retrospective cohort study of healthcare workers who had self-isolated due to COVID-19.

**Setting:** University Hospitals Birmingham NHS Foundation Trust, UK (UHBFT).

**Participants:** 956 health care workers were recruited by open invitation via UHBFT trust email and social media.

**Intervention:** Participants volunteered a venous blood sample that was tested for the presence of anti-SARS-CoV-2 spike glycoprotein antibodies. Results were interpreted in the context of the symptoms of their original illness and ethnodemographic variables.

**Results:** Using an assay that simultaneously measures the combined IgG, IgA and IgM response against the spike glycoprotein (IgGAM), the overall seroprevalence within this cohort was 46.2% (n=442/956). The seroprevalence of immunoglobulin isotypes was 36.3%, 18.7% and 8.1% for IgG, IgA and IgM respectively. IgGAM identified serological responses in 40.6% (n=52/128) of symptomatic individuals who reported a negative SARS-CoV-2 PCR test. Increasing age, non-white ethnicity and obesity were independently associated with greater IgG antibody response against the spike glycoprotein. Self-reported fever and fatigue were associated with greater IgG and IgA responses against the spike glycoprotein. The combination of fever and/or cough and/or anosmia had a positive predictive value of 92.3% for seropositivity.

**Conclusions and relevance:** Assays employing combined antibody detection demonstrate enhanced seroepidemiological sensitivity and can detect prior viral exposure even when PCR swabs have been negative. We demonstrate an association between known ethnodemographic risk factors associated with mortality from COVID-19 and the magnitude of serological responses in mild-to-moderate disease. The combination of cough, and/or fever and/or anosmia identifies the majority of individuals who should self-isolate for COVID-19.

## Introduction

In the general population, increasing age, male sex, obesity, non-white ethnicity, socioeconomic deprivation and co-morbidities leading to direct or indirect immune suppression are established risk factors associated with mortality from COVID-19 [1]. In hospitalised patients, severe COVID-19 is associated with peripheral blood signatures suggestive of dysregulated interferon responses, T cell exhaustion and high antibody production [2-5]. Whether high-risk ethnodemographic variables are directly associated with dysregulated immunological responses in severe COVID-19 is not known. Furthermore, whether ethnodemographic variables are associated with differential serological responses against SARS-CoV-2 in mild disease is also unknown.

Healthcare workers provide a unique cohort in which to consider the underlying immunology of SARS-CoV-2 infection. Healthcare workers are at high risk of exposure to SARS-CoV-2 during the course of their work; estimates of infection rates and seroprevalence in cohorts of UK healthcare workers consistently exceed those of the general population [6-8]. Furthermore, cohorts of healthcare workers tend to be young, ethnically diverse and less co-morbid compared to hospitalised patients.

In this study, using a cohort of UK healthcare workers, we define the serological response directed against the SARS-CoV-2 spike glycoprotein of non-hospitalised adults following mild or moderate COVID-19 and explore the relationships between that serological response and ethnodemographic variables that are associated with poor outcome from COVID-19. We also explore associations between disease symptomatology and the serological response. Finally, we consider the cumulative occupational risk faced by UK healthcare workers over the course of the first wave of the COVID-19 and the impact of self-isolation periods on healthcare delivery.

## Methods

A cohort of healthcare workers who had previously self-isolated because they experienced symptoms suggestive of COVID-19, or self-isolated because household contacts had experienced symptoms of COVID-19 were recruited to this study between 27/4/2020 and the 08/06/2020. Open invitation to the study was made via UHBFT email to all staff and also advertised via social media. The only pre-defined exclusion criteria was participation in an existing SARS-CoV-2 vaccine trial or current COVID-19 symptomatology. No individuals within this cohort were hospitalised with COVID-19.

All individuals volunteered a venous blood sample that was tested for anti-SARS-CoV-2 spike glycoprotein antibodies using a commercially available IgGAM ELISA that measures the total antibody response (Product code: MK654, The Binding Site (TBS), Birmingham). The SARS-CoV-2 spike utilized in the ELISA is a soluble, stabilized, trimeric glycoprotein truncated at the transmembrane region [9, 10]. This assay has been CE-marked with 98.3% (95% CI: 96.4-99.4%) specificity and 98.6% sensitivity (95% CI: 92.6-100%) following PCR proven, non-hospitalised, mild-to-moderate COVID-19. Further serological investigations were undertaken in individuals who were found to be seropositive on this screening assay. TBS anti-SARS-CoV-2 spike plates were also used to assess individual IgG, IgA, and IgM antibodies. Serum was pre-diluted at a 1:40 dilution using a Dynex Revelation automated liquid handler (Dynex, USA). Antibodies were detected using sheep-anti-human HRP-conjugated polyclonal antibodies against IgG (1:16,000), IgA (1:2000), and IgM (1:8000) (TBS, UK). Plates were developed after 10 minutes using TMB core (TBS, UK), and orthophosphoric acid (TBS, UK) used as a stop solution. Optical densities at 450_nm_ (OD_450nm_) were measured using the Dynex Revelation automated liquid handler. IgG, IgA, and IgM ratio-cutoffs were determined based on running 90 pre-2019 negative serum samples. A cutoff ratio relative to the TBS cutoff calibrators was determined by plotting the pre-2019 negatives (n=90) in a frequency histogram chart. Once the ratio cutoff was determined from the pre-2019 negatives, a cut-off coefficient was established for IgG (1), IgA (0.71), and IgM (0.588). Any ratio values > 1, are classed as positive. Any ratio values < 1 are classed as negative.

At enrolment the following variables were recorded: age, sex, ethnicity, height and weight, number of co-occupants in participants household, whether an individual used public transport in the two weeks prior to their isolation period, the dates of their isolation period, their job role, the department in which they worked during the months of March 2020 to June 2020, whether they had undergone a previous PCR test for SARS-CoV-2 and the result of that test. Participants were also asked to retrospectively report whether, during their acute illness for which they self-isolated, they suffered any of the following symptoms: cough, shortness of breath, sore throat, fever > 37.8°C, fatigue, myalgia, anosmia and diarrhoea. UHBFT inpatient data was sourced by the UHBFT infection control team. The index of multiple deprivation rank from participants home postcodes were sourced from 2019 UK Ministry of Housing, Communities and Local Government statistics [11] and transformed into a normally distributed score using the function [log(R/(32,844-R)] where R represented the individual rank of a participant’s postcode within the national data.

Data were analysed using Graph Pad Prism 9.0. Categorical data was compared using the Chi squared test and optical density distributions using the Kruskal-Wallis test with Dunn’s post-test comparison for individual groups. Seroprevalence data are expressed as a percentage, with binomial confidence intervals calculated using Wilson’s method. The relationship between age, body mass index (BMI), and antibody responses was considered using Pearson’s correlation coefficient. The relationship between antibody levels and time from symptom onset was modelled using a smoothing spline curve with 4 knots. Multiple logistic regression was performed using seropositivity as the outcome variable. Age, sex, ethnicity, household index of multiple deprivation score, household occupants, whether an individual experienced primary symptoms or isolated due to a household contact becoming unwell and public transport use were included as independent variables. For continuous variables, the odds ratio represents change in odds of seropositivity per unit increase the independent variable. Multiple linear regression was performed using the IgG, IgA and IgM ratios as outcome variables and age, sex, ethnicity, BMI, household index of multiple deprivation score and time from symptom onset as independent variables.

The study was approved by the London - Camden & Kings Cross Research Ethics Committee reference 20/HRA/1817. All participants provided written, informed consent prior to enrolment in the study.

## Results

Nine hundred and fifty-six healthcare workers were enrolled in this study (**Table 1**). Using the combined anti-IgG, IgA and IgM (IgGAM) antibody assay, the overall seroprevalence of anti-SARS-CoV-2 antibodies in the cohort was 46.2% (n=442/956) (**Figure 1A**). Age, sex, number of household co-occupants, public transport use and index of multiple deprivation scores associated with participants home postcodes did not significantly influence seroprevalence (**Table 1, Supplementary Figure 1**). However, ethnicity did have an effect with individuals of Black (72.2% seropositive 95% CI 56.0 - 84.2%) and Asian ethnicity (54.1% seropositive, 95% CI 46.2 -61.4%) demonstrating the highest seroprevalence (overall Chi square 19.2, degrees of freedom 5, p=0.002) (**Supplementary Figure 1**). Individuals who self-isolated because a household contact had experienced symptoms suggestive of SARS-CoV-2 infection (n=162/423) were significantly less likely to be seropositive at the time of the study than those who self-isolated because they experienced symptoms directly (n = 243/467) (38.3% vs 52.0%, Chi-square=16.89, z=4.11, df=1, p<0.0001). When these variables were considered in a multiple logistic regression model (**Table 2**), Black, Asian and minority ethnic (BAME) ethnicity (OR 1.90 (95% CI 1.30-2.81), z=3.26, p=0.001) was the only statistically significant risk factor for seropositivity.

**Table 1:** Demographics of study population. Median and interquartile ranges are provided. Age was compared using a two-tailed unpaired Mann-Whitney test. Categorical data was compared using the Chi Square test. The index of multiple deprivation scores were compared using an unpaired two-tailed T test.

|  | All participants,<br>n (%) | Seropositive<br>n (%) | Seronegative<br>n (%) | Seroprevalence<br>(%) | P value |
| --- | --- | --- | --- | --- | --- |
| n | 956 | 442 | 514 | 46.2% |  |
| Age (years) | 41.0<br>(31.0-50.0) | 41.0<br>(32.0-50.0) | 40.0<br>(31.0-50.0) | - | 0.69 (MW) |
| Sex |  |  |  |  | 0.33 (ChiS) |
| -Male | 260<br>(27.2%) | 110<br>(24.9%) | 150<br>(29.2%) | 42.6% |  |
| -Female | 679<br>(71.0%) | 324<br>(73.3%) | 355<br>(69.1%) | 47.7% |  |
| - Not stated | 17<br>(1.8%) | 8<br>(1.8%) | 9<br>(1.8%) | 47.0% |  |
| Ethnicity |  |  |  |  | 0.002 (ChiS) |
| - White | 691<br>(72.3%) | 294<br>(66.5%) | 397<br>(77.2%) | 42.5% |  |
| - Mixed | 22<br>(2.3%) | 10<br>(2.3%) | 12<br>(2.3%) | 45.5% |  |
| - Asian | 170<br>(17.8%) | 92<br>(20.8%) | 78<br>(15.2%) | 54.1% |  |
| - Black | 36<br>(3.8%) | 26<br>(5.9%) | 10<br>(1.9%) | 72.2% |  |
| - Other | 25<br>(2.6%) | 13<br>(2.9%) | 12<br>(2.3%) | 52.0% |  |
| - Not stated | 12<br>(1.3%) | 7<br>(1.6%) | 5<br>(1.0%) | 58.3% |  |
| Index of multiple<br>deprivation score | 780 | -0.04 (0.82) | -0.04 (0.77) |  | 0.99 (t-test) |

**Table 2:**
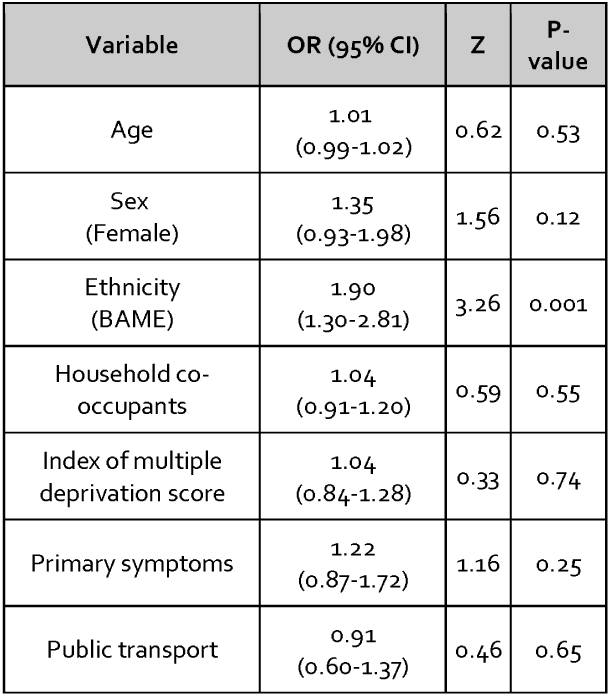
Multiple logistic regression of factors affecting seropositivity. Seropositivity at the time of study enrolment was used as the dependent variable. Participants’ age, sex, ethnicity (white vs BAME), number of household co-occupants, the index of multiple deprivation score, whether an individual isolated because they directly experienced symptoms or isolated because a family member experienced symptoms and public transport use in the two weeks prior to isolation were used as independent variables. Odds ratios (OR) and 95% confidence intervals (CI) are provided. The area under the receiver operator curve of this model was 0.58, p=0.0007.

**Figure 1:**
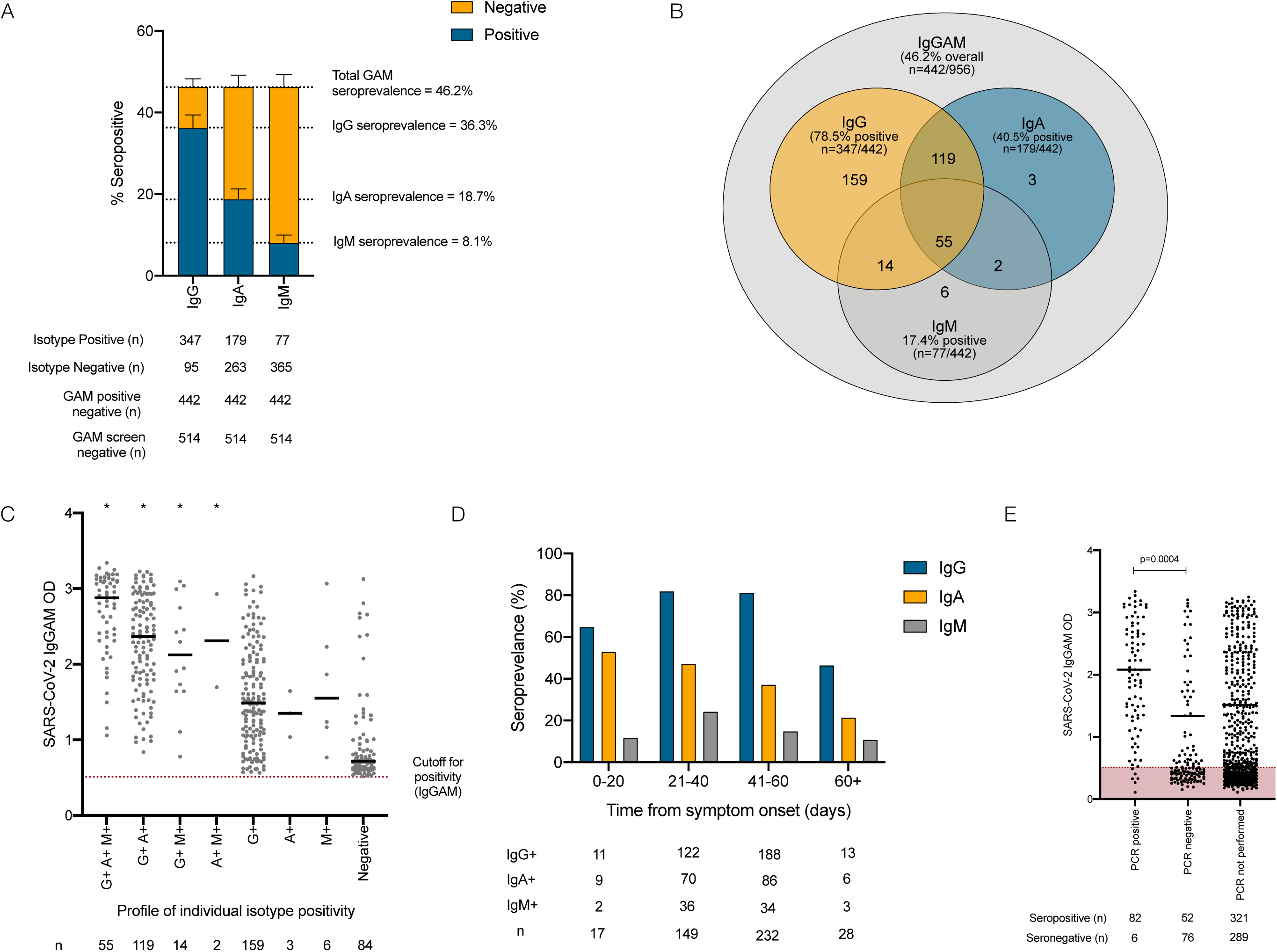
Serological response against the SARS-CoV-2 spike glycoprotein in healthcare workers. (**A**) IgG, IgA and IgM responses in individuals demonstrating seropositivity in the combined IgGAM ELISA. (**B**) Venn diagram illustrating the relationship between IgG, IgA and IgM seropositivity in this cohort. (**C**) Optical densities (OD) of the total serum antibody response determined by the combined IgGAM assay, in individuals with different patterns of IgG, IgA and IgM isotype seropositivity. Bars represent the median of all results above the assay cutoff. * represents p<0.0001 (Kruskal-Wallis, Dunn’s post-test comparison) of each group compared to the group only detectable using the IgGAM assay (**D**) Seroprevalence of IgG, IgA and IgM isotypes in relation to time from symptom onset. (**E**) Optical densities (OD) of the total serum antibody response determined by the combined IgGAM assay in symptomatic individuals who had previously undergone PCR testing for the SARS-CoV-2. Bars represent the median of all results above the assay cutoff.

The 442 seropositive individuals had their antibody response characterised further by measuring the individual immunoglobulin isotypes (IgG, IgA and IgM) against the viral spike glycoprotein (**Figure 1A**). IgG antibodies were detectable in 36.3% (n=347/956), IgA antibodies 18.7% (n=179/956) and IgM antibodies 8.1% (n=77/956). The combined IgGAM assay identified 9.9% (n=95/956) of participants who demonstrated a serological response against the viral spike glycoprotein that would not have been detected if IgG detection alone was used in an equivalent assay (**Figure 1B**). The enhanced analytical sensitivity of combined IgGAM detection arises from the identification of seropositivity in individuals who fall below the limit of detection of the equivalent assays that measure individual immunoglobulin isotypes in isolation (**Figure 1C**). Of the 347 individuals who were seropositive for IgG, 50.1% (n=174/347) also demonstrated IgA antibodies in the serum and 19.8% (n=69/347) demonstrated IgM antibodies in the serum (**Figure 1B**). Exclusive IgA or IgM seropositivity was rare (n=3 for IgA, n=6 for IgM). The enhanced sensitivity demonstrated by combined IgGAM detection may facilitate the identification of seropositive individuals beyond 60 days from symptom onset, where detectable IgG seropositivity falls to 46.4% (n=13/28) (**Figure 1D, Supplementary Figure 2**). In this study, only 26.6% (n=216/812) of symptomatic participants received confirmatory PCR testing reflecting the lack of access to community testing at the time. The IgGAM assay identified 93.2% (n=82/88) of individuals who had previously tested positive for SARS-CoV-2 by PCR and an additional 52 previously symptomatic individuals who had tested negative by PCR; these individuals had significantly lower antibody levels than those who had tested positive by PCR (**Figure 1E**). Differences in the magnitude of the total antibody response against the spike glycoprotein between PCR positive and PCR negative participants could not be explained by differences in the time allowed for the maturation of the antibody response, which was equivalent between the groups (time from symptom onset: 33.8 days vs. 37.1 days, p=0.18).

**Figure 2:**
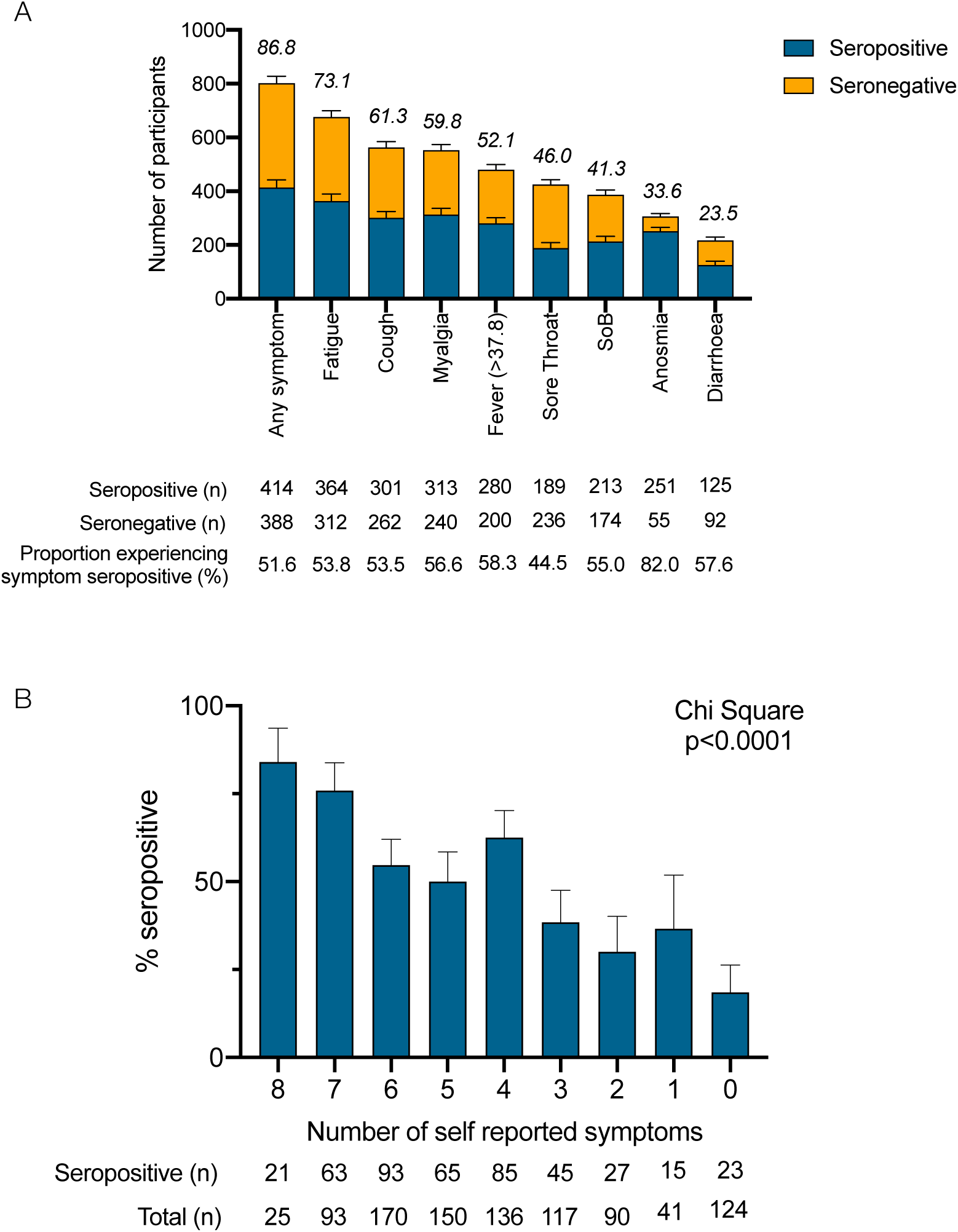
Self-reported symptoms in relation to seropositivity in healthcare workers. (**A**) Self-reported symptoms in relation to seropositivity in healthcare workers. Numbers above bars represent the percentage of participants experiencing symptom. (**B**) Number of self-reported symptoms in relation to seropositivity in healthcare workers; data was compared using Chi-square test (Chi Square = 114.8, df =8, p<0.0001).

If previously symptomatic, participants were asked to describe their main symptoms of COVID-19 at study enrolment (**Table 3, Figure 2A**). 86.8% (n=812/926) experienced at least one symptom they attributed to SARS-CoV-2 infection. Individuals who tested negative by PCR, self-reported fewer symptoms than those testing positive by PCR (average number of symptoms: 4.6 vs. 5.1, p=0.02). Fatigue was the most common symptom, experienced by 73.1% (n=676/925) of participants and demonstrated the highest specificity for seropositivity at 71.1%. Anosmia was the most sensitive symptom (82.0%) in relation to overall seropositivity but was only reported by 33.5% (n=306/911) of participants. The combination of cough and/or fever and/or anosmia was experienced by 78.6% of participants and captured 92.3% of individuals who were seropositive at the time of the study enrolment. The likelihood of an individual testing positive for SARS-CoV-2 antibodies progressively increased with the number of self-reported symptoms (Chi square 129.9, degrees of freedom = 16, p<0.0001) (**Figure 2B**).

**Table 3:** Performance characteristics of self-reported symptoms in relation to seropositivity at study enrolment.

| Symptom | Number of participants experiencing symptom (n) | Participants experiencing symptoms (%) | Sensitivity (%) | Specificity (%) | Positive predictive value (%) | Negative predictive value (%) |
| --- | --- | --- | --- | --- | --- | --- |
| Fatigue | 676 | 73.1 | 53.8 | 71.1 | 83.5 | 36.2 |
| Cough | 563 | 61.3 | 53.5 | 62.9 | 69.5 | 46.1 |
| Myalgia | 553 | 59.8 | 56.6 | 65.4 | 71.6 | 49.4 |
| Fever > 37.8°C | 480 | 52.1 | 58.3 | 64.7 | 64.2 | 58.8 |
| Sore Throat | 425 | 46.0 | 44.5 | 50.9 | 43.5 | 51.8 |
| Shortness of Breath | 387 | 41.3 | 55.0 | 59.1 | 48.6 | 65.1 |
| Anosmia | 306 | 33.6 | 82.0 | 70.4 | 58.4 | 88.6 |
| Diarrhoea | 217 | 23.5 | 57.6 | 55.8 | 28.6 | 81.1 |
| Cough *or* Fever *or* Anosmia | 752 | 78.6 | 54.3 | 83.3 | 92.3 | 33.1 |

The relationship between symptoms and the magnitude of IgG, IgA and IgM responses directed against the SARS-CoV-2 spike glycoprotein was analysed (**Supplementary Figure 3**). Self-reported fever and fatigue were associated with significantly greater IgG and IgA responses against the viral spike glycoprotein, while self-reported diarrhoea was associated with significantly greater IgG responses. These symptoms may be associated with a greater degree of systemic illness arising from SARS-CoV-2 infection.

**Figure 3:**
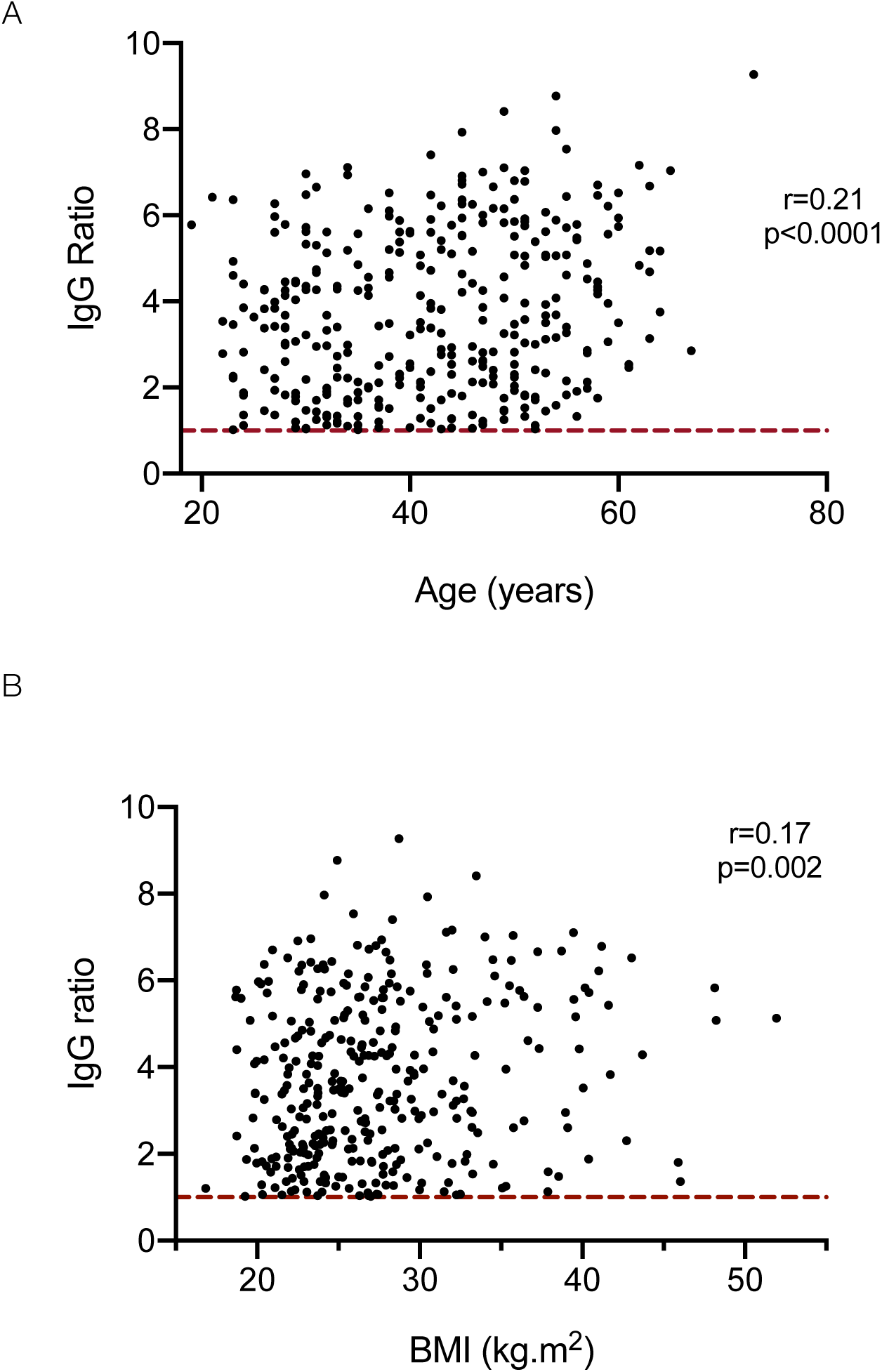
Relationship between age (A) and body mass index (B) and the magnitude of the IgG response against the SARS-CoV-2 spike glycoprotein. Dotted red line represents assay cut-off.

The relationship between ethnodemographic variables and the magnitude of IgG, IgA and IgM responses was analysed. Sex did not significantly affect the magnitude of response of any antibody isotype (**Supplementary Figure 4A**). However, increasing age was associated with a higher IgG response against the viral spike glycoprotein (**Supplementary Figure 4B**): a weak, but statistically significant positive correlation was observed between age and the magnitude of the IgG response (Pearson correlation r=0.21, p<0.0001) (**Figure 3A**) and when analysed by age brackets, the median IgG response in individuals aged 56-65 was significantly higher than those aged 26-35 (Kruskal-Wallis statistic 14.0, p=0.02, Dunn’s post-test comparison between 26-35 vs. 56-65 year old age groups; mean rank difference -65.22, p=0.01) (**Supplementary Figure 4B**). Individuals from all non-white ethnic groups demonstrated higher median antibody levels than white individuals with significantly greater levels observed in Asian individuals compared to white individuals (Kruskal-Wallis statistic 16.9, p=0.005, Dunn’s post-test comparison between white vs. Asian ethnic groups; mean rank difference -37.55, p=0.03) (**Supplementary Figure 4C**). Increasing BMI was also associated with increased IgG responses against the viral spike glycoprotein (Kruskal-Wallis statistic 12.1, p=0.03) with a weak but significant correlation (r=0.17, p=0.002) (**Figure 3B, Supplementary Figure 4D**). A linear regression model incorporating these variables demonstrated increasing age, non-white ethnicity, and increasing BMI were independently associated with greater IgG responses and non-white ethnicity significantly associated with greater IgM responses (**Table 4**).

**Table 4:**
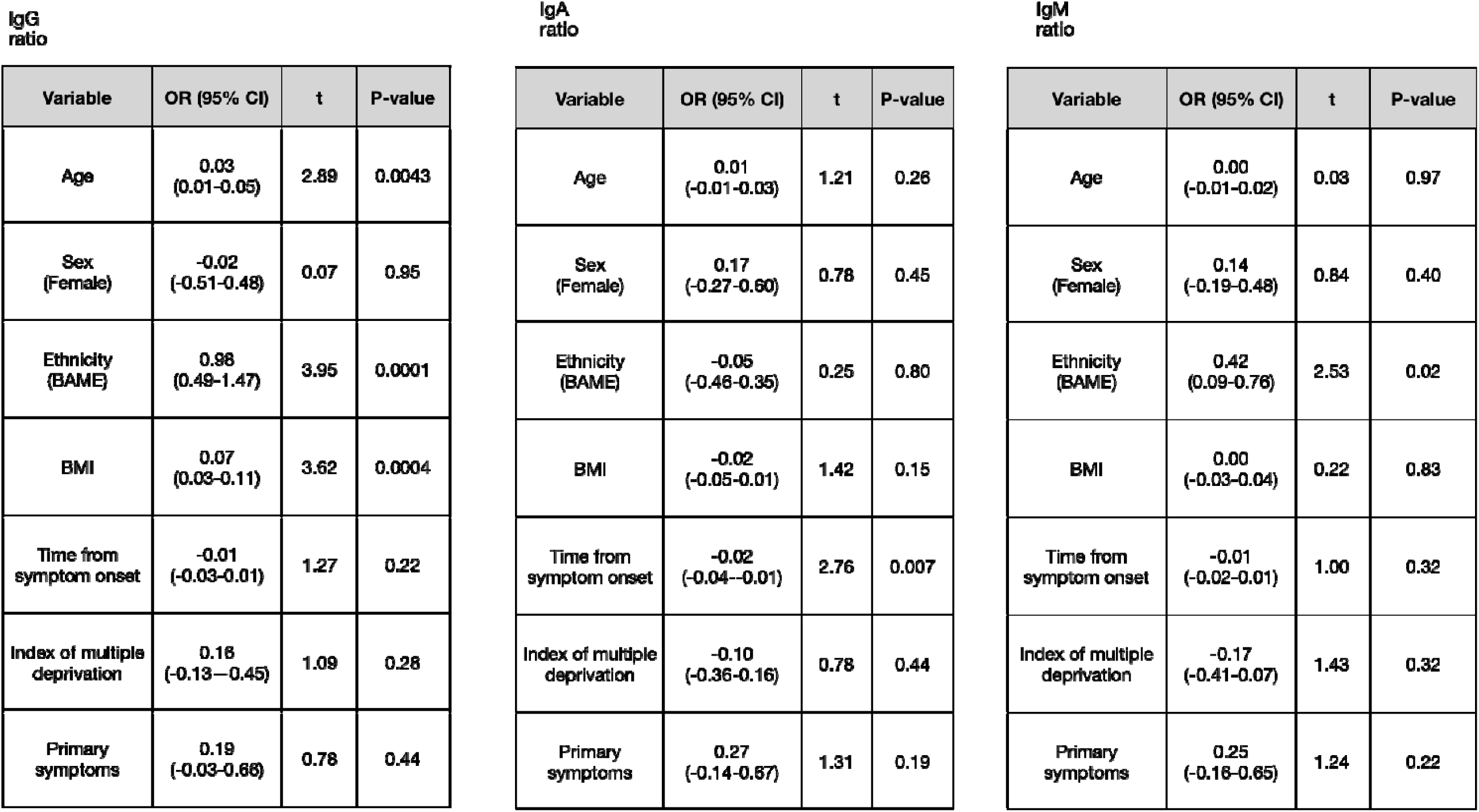
Linear regression models of variables affects the magnitude of the antibody response against the SARS-CoV-2 spike glycoprotein. The IgG, IgA and IgM ratios were used as dependent variables and participants’ age, sex, ethnicity, body mass index, time from symptom onset, the index of multiple deprivation score, whether an individual isolated because they directly experienced symptoms or isolated because a family member experienced symptoms and public transport use in the two weeks prior to isolation were used as independent variables. Odds ratios (OR) and 95% confidence intervals (CI) are provided. For continuous variables, the OR represents the increase in immunoglobulin ratio associated with each unit increase in that variable. For categorical variables, the OR represents the increase in immunoglobulin ratio associated the variable in parenthesis.

**Figure 4:**
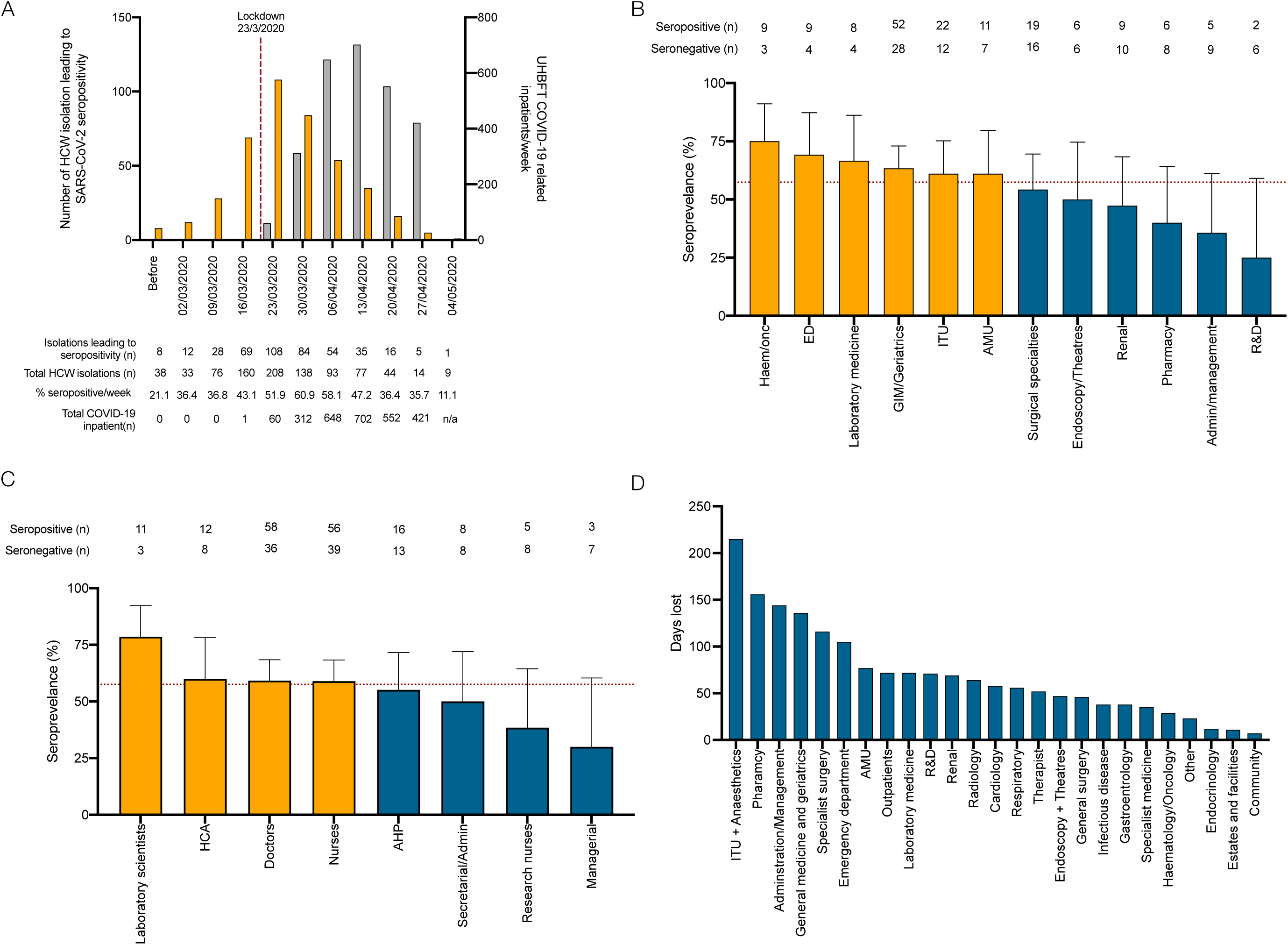
COVID-19 risk in healthcare workers. (A) Timing of isolation events in study participants, seroconversion rates and UHBFT COVID-19 positive inpatients from February to May 2020. (B) Hospital departments and job roles (C) of participants who self-isolated because they directly experienced symptoms following the arrival for the first COVID-19 inpatient at UHBFT. (D) Number of potential days lost due to isolation events in individuals who did not have a PCR test and were found to be seronegative at study enrolment.

With respect to the timing of infections and occupational risk of exposure in healthcare workers, the proportion of self-isolations associated with seropositivity at the time of study enrolment progressively increased from 21.1% (n=8/38) in February 2020 to a peak of 60.9% (n=84/138) in the week beginning 30th March 2020 before declining during April and May 2020 (**Figure 4A**). By the time of UK national lockdown (23^rd^ March 2020), when only 60 proven COVID-19 patients had been admitted to UHBFT, 53.6% (n=225/420) of self-isolations associated with seropositivity had already occurred. By exclusively considering individuals who had isolated after 23^rd^ March 2020, the occupational risk of healthcare workers was reconsidered (**Figure 4B and 4C**). Seroprevalence in this selected cohort was 57.5% (n=176/306); data was mapped to job roles and hospital departments. Seroprevalence was greater in departments that were directly patient facing (haematology/oncology (75.0%), emergency department (69.2%), general medicine and geriatrics (63.4%)) and lower in non-patient facing roles (administration/management (35.7%), research and development (25.0%). Laboratory scientists had the highest seroprevalence of any healthcare worker group in this study (78.6%); healthcare assistants, doctors, nurses and allied healthcare professionals all had similar seroprevalence (55.2-60.0%). Multiple logistic regression in this subgroup demonstrated no particular department or job role was at significantly greater risk of seropositivity; however, non-white ethnicity significantly increased the risk of seropositivity in models considering job role and department (**Supplementary Table 1**). Assuming individuals who were seronegative at the time of enrolment in the study were unexposed to the virus, we estimate a total of 1749 working days were lost due to healthcare workers isolating for symptoms that were not attributable to the virus, representing 16.4% of the total working days lost (n=1749/10670). ITU and anaesthetics experienced the greatest burden with a total of 215 working days lost (**Figure 4D**).

## Discussion

Severe COVID-19 is associated with immune dysregulation, multi-organ dysfunction and death. Age, obesity and non-white ethnicity have been independently associated with poor outcome from COVID-19 [1, 12]. In this study, we demonstrate that these risk factors are independently associated with greater IgG responses directed against the SARS-CoV-2 spike glycoprotein. Exaggerated serological responses have previously been observed in severe COVID-19 [13, 14], however, by conducting this study in individuals with mild disease, it is unlikely our findings are non-specific artefacts of prolonged critical illness. Instead, discreet pathogenic mechanisms are likely to be associated with each variable that require further delineation.

Increasing age is associated with immunosenescence, a phenomenon characterised by complex and progressive immunological changes resulting in increased susceptibility to infectious disease [15]. As response to vaccination diminishes with age [16], it was not anticipated that increasing age would be associated with greater SARS-CoV-2 IgG antibodies. However, this cohort only included individuals of working age and further studies are required to see whether this effect persists in older age groups. Investigation should also consider the quality of the antibody response; for example, increasing age is associated with poor functionality of anti-pneumococcal antibodies and discordance has been noted between absolute antibody titres and functionality in the co-morbid elderly [17, 18].

Obesity has been postulated to increase mortality from COVID-19 by reducing physiological cardio-respiratory reserve and facilitating a pro-thrombotic state [19]. Whether obesity directly affects immunological responses is less clear. Adipose tissue is known to release interleukin-6 [20] which indirectly induces B lymphocyte antibody production via T lymphocyte derived interleukin-21 [21]. Furthermore, increased BMI is associated with low-grade systemic inflammation evidenced by increased serum C-reactive protein, an acute phase protein which is IL-6 dependent [22, 23]. Further studies exploring the relationship between obesity, adiposity, baseline IL-6 levels and the magnitude and quality of antiviral antibody responses may facilitate enhanced patient selection when considering the use of IL-6 blockade in COVID-19 infection [24].

Non-white ethnicity is associated with poorer outcomes from COVID-19 [1]. It is also associated with either an increased risk of infection from SARS-CoV-2, or an increased proportion of infections that drive serologically detectable antiviral antibody response [6, 8, 25]. Socioeconomic differences leading to increased viral exposure have been postulated to account for these differences [26], but in this study, household occupancy and deprivation scores associated with a participant’s home postcode were not associated with SARS-CoV-2 seropositivity. BAME ethnicity was, however, independently associated with greater IgG and IgM directed against the SARS-CoV-2 spike glycoprotein. The peripheral immunophenotypes of healthy individuals differs by ethnicity [27]: individuals of African American ethnicity have significantly greater proportions of type 17 T-follicular helper cells, significantly lower type 1 T follicular helper cells, significantly higher proportions of B cells within their peripheral lymphocyte populations and higher levels of immunoglobulins in comparison to white individuals [27, 28]. Whether an individual’s peripheral immunophenotype correlates with acute antibody responses to SARS-CoV-2 is not known. The epidemiology and genomic architecture underlying differential ethnic susceptibility to antibody-mediated diseases may provide insight into the immune response against COVID-19 [29]. Equally, expression of the ACE-2 receptor, necessary for viral entry, may vary between sex and ethnic groups leading to differential risk of infection upon viral exposure.

Our study has implications for future SARS-CoV-2 seroprevalence studies. Previously, we have demonstrated the superior sensitivity of the trimeric, native-like SARS-CoV-2 spike glycoprotein in comparison to the nucleocapsid for the detection of antibody responses in individuals with mild COVID-19 [30]. We now demonstrate that measuring the total antibody response directed against the SARS-CoV-2 spike glycoprotein, is more sensitive than measuring an individual immunoglobulin isotype in isolation. It has been postulated that SARS-CoV-2 seroprevalence may be underestimated by not considering systemic IgA responses against virus [31]. We unequivocally demonstrate that a minority of individuals exclusively mount IgA responses and its independent measurement is unlikely to significantly affect estimates of seroprevalence. However, a combined approach that measures the total antibody responses greatly enhances assay sensitivity in mild disease and should be considered in future seroprevalence studies. Furthermore, these data highlight potential limitations in PCR testing to confirm acute COVID-19. Only 26.6% of symptomatic individuals received a PCR test highlighting the lack of available testing during the first-wave of the COVID-19 pandemic. However, an antibody response was detectable in 40.6% of symptomatic individuals who tested negative by PCR, although the magnitude of this response was significantly less than those who tested PCR positive. This was not explained by differences in the time allowed for maturation of the antibody response which was equivalent between the groups, but notably, patients who tested PCR negative reported, on average, fewer symptoms than those who tested PCR positive. Previous studies have demonstrated the upper respiratory tract viral load, estimated by PCR cycle threshold values, is equivalent in asymptomatic and symptomatic individuals [32]. These data would support a hypothesis that some individuals may experience fewer symptoms because they achieve more rapid immunological control over viral replication; this in turn may narrow the window of PCR positivity and highlight potential end-to-end operational insensitivities when PCR is used for the detection of mild disease. Such issues have previously been highlighted in more seriously unwell hospitalised patients [33, 34] and must be very carefully considered when PCR is used as the gold-standard diagnostic reference point to assess the performance of other molecular and serological assays.

With respect to the sustainable delivery of healthcare during future pandemic infections, this study contributes a number of important observations: firstly, the overall seroprevalence within this cohort, selected because they had self-isolated was 46.2%. 27.8% of illnesses leading to seroconversion in healthcare workers occurred prior to the arrival of PCR confirmed COVID-19 patients within the hospital environment and 53.6% of illnesses leading to seroconversion had occurred by the end of the following week. Given the median incubation time of the virus is 5 days, these data strongly suggest that the majority of COVID-19 in hospital-based healthcare workers was not acquired from known COVID-19 inpatients in this wave. It also raises the possibility of pre-symptomatic healthcare workers introducing SARS-CoV-2 into the hospital environment; our previous study demonstrated 2.4% of asymptomatic healthcare workers tested positive for SARS-CoV-2 nucleic acid on nasopharyngeal swabs while at work [8].

Nevertheless, relatively increased seroprevalence was observed in direct patient facing workgroups, in comparison to those with minimal or no patient contact suggesting an occupational risk of exposure to SARS-CoV-2 exists. That risk was homogenous for all patient facing groups (55.2%-60.0% seroprevalence in healthcare assistants, doctors, nurses and allied health professional). Of note, laboratory scientists had exceptionally high seroprevalence, possibly due to recirculation of aerosolised virus within temperature-controlled laboratories [35]. A considerable number of working days were lost to staff members isolating for symptoms that were not molecularly or serologically proven to be COVID-19. It is reassuring that the combination of cough or fever or anosmia captures 92.3% of individuals who tested seropositive at study enrolment, validating these symptoms as an effective, but not perfect, way of determining who should self-isolate. However, by understanding the performance characteristics of molecular and serological assays in respect to infection and infectivity following SARS-CoV-2, medical human resources may be better managed.

Our study is benefited by the large cohort enrolled but limited by its retrospective nature, its focus on individual of working age and that individuals were asked to self-report symptoms. By selecting self-isolating individuals, the cohort is enriched for individuals who will have had COVID-19: while this allows the study of factors affecting the magnitude of the antibody response, it excludes individuals who may have been asymptomatic and who may not mount an antibody response. Nevertheless, the variables we identify as affecting the antibody response are known population level risk factors for poor outcome and it is plausible an immunological mechanism is implicated in disease pathogenesis. Further studies must continue to explore these associations, particularly in mild disease, to inform COVID-19 pathogenesis.

## Data Availability

The anonymised dataset will be made available on reasonable request. Proposals should be directed to the corresponding author.

## Supplementary Figure Legends

**Supplementary Figure 1:** Seropositivity in healthcare workers by: (A) age, (B) sex, (C) ethnicity,

(D) number of household co-occupants, (E) Public transport use in the two weeks prior to isolation period.

**Supplementary Figure 2:** Anti SARS-CoV-2 spike glycoprotein antibody kinetics over time as detected using: (A) IgGAM assay, (B) IgG assay, (C) IgA assay, (D) IgM assay. Red area represents assay cut-off. Spline modelling has been used to model change over time.

**Supplementary Figure 3:** IgG, IgA and IgM anti SARS-CoV-2 spike glycoprotein antibody responses in relation to participants ‘self-reported symptoms. Horizontal black lines represent the median of positive results in each group. Red areas represent ratios below the assay cut-off. Medians of positive results were compared using two tailed Mann-Whitney test.

**Supplementary Figure 4:** IgG, IgA and IgM anti SARS-CoV-2 spike glycoprotein antibody responses in relation to: (A) sex, (B) age, (C) ethnicity, (D) class of obesity. Horizontal black lines represent the median of positive results in each. The medians from each group were compared using the Kruskal-Wallis test; statistically significant values represent Dunn’s post-test comparison.

## Acknowledgments

The authors would like to acknowledge the staff of the Clinical Immunology Service who helped process the samples for serological testing, Dr Margaret Goodall for her expertise in antibody production and assay development, Dr Jason McLellan for the expression plasmid for the SARS-CoV-2 glycoprotein. We would also like to acknowledge all the participants from UHBFT. Serological assay development was undertaken in collaboration with The Binding Site Group Ltd.

## Author Contributions

AMS and SEF helped conceive the study, performed experiments, collated and analysed the data, produced the figures, wrote and revised the manuscript and should be considered joint first authors. MPT and SJ performed experiments, collated and analysed the data. JDA, YW and MC produced the original trimeric spike-glycoprotein on which the serological assays are based and advised on methodology. AG, GM and JON recruited participants to the study, facilitated the acquisition of clinical samples and collated study results. MIG collated and interpreted trust-level data on infections within UHBFT inpatients. SAT, CB, LD, DE, BE and AF processed samples, undertook experiments and collated results for serological studies. MIG provided trust inpatient data. DCW, AFC and MTD helped conceive the study and supervised analysis of data from the study. AGR is the senior and corresponding author for this manuscript and provided overall leadership for all aspects of the study. All authors helped revise the manuscript for publication.

