## Supplementary figures and images for "Serological responses to SARS-CoV-2 following non-hospitalised infection: clinical and ethnodemographic features associated with the magnitude of the antibody response"

### Supplementary Figures

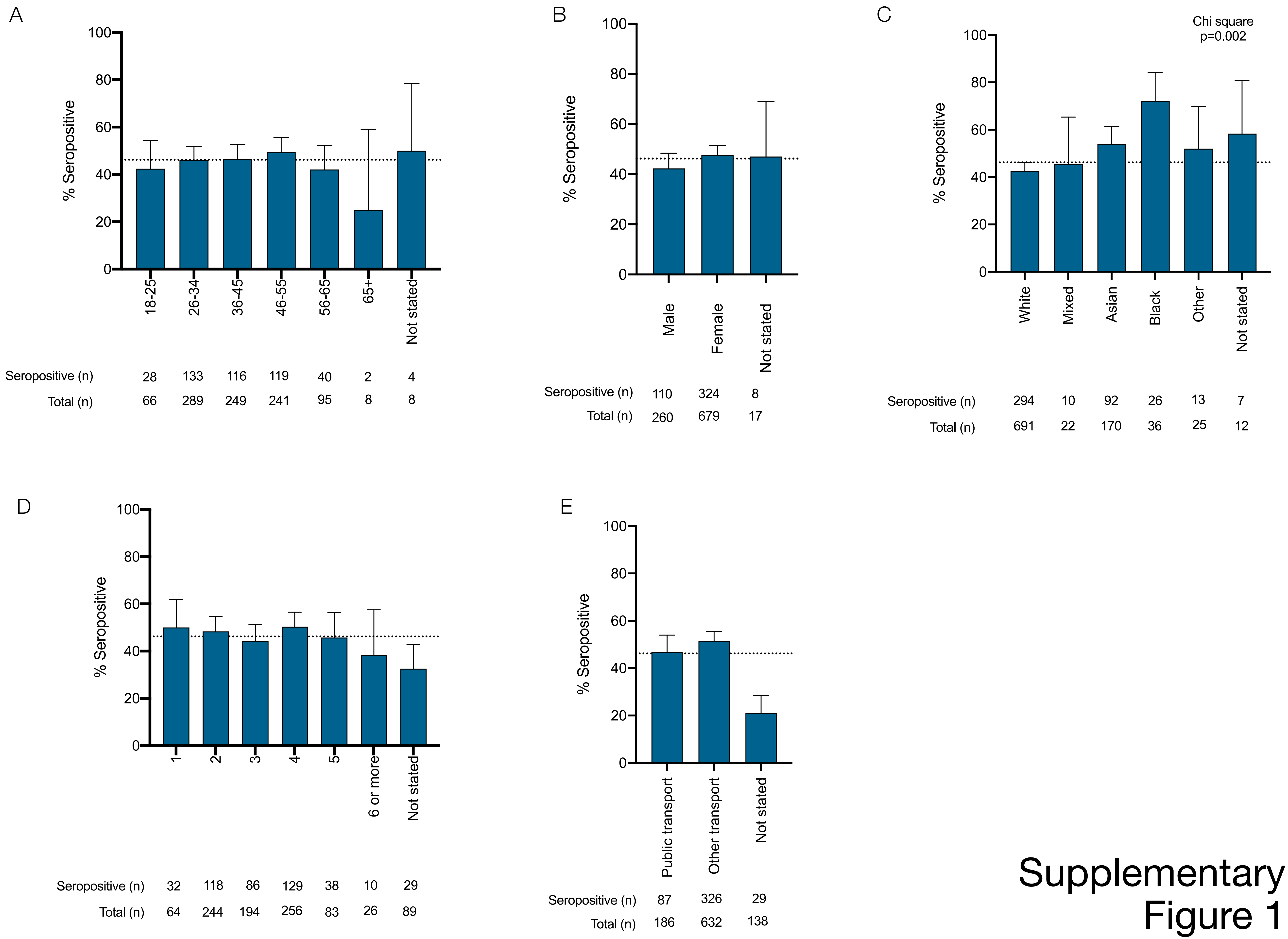


Supplementary Figure 1

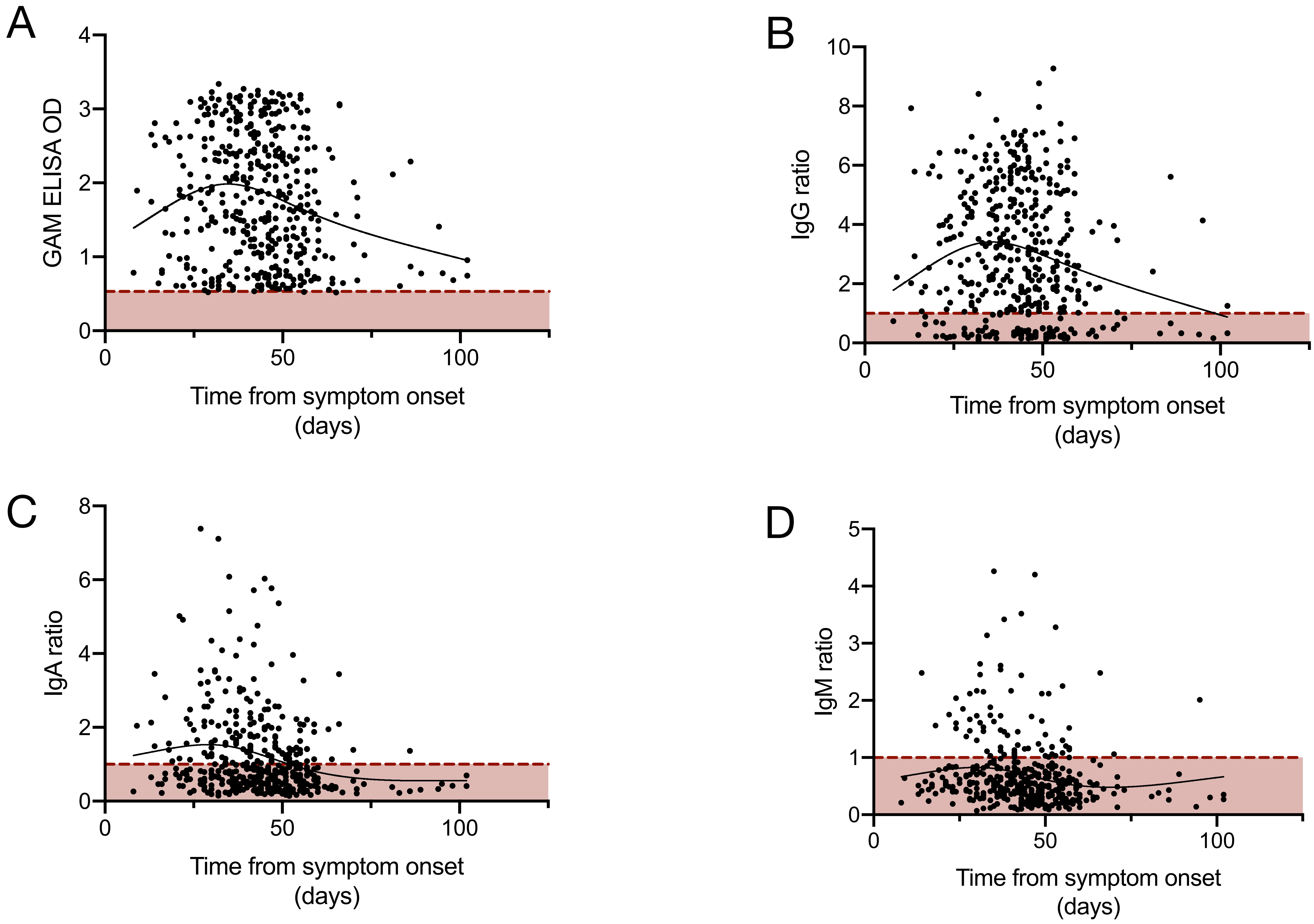


Supplementary Figure 2

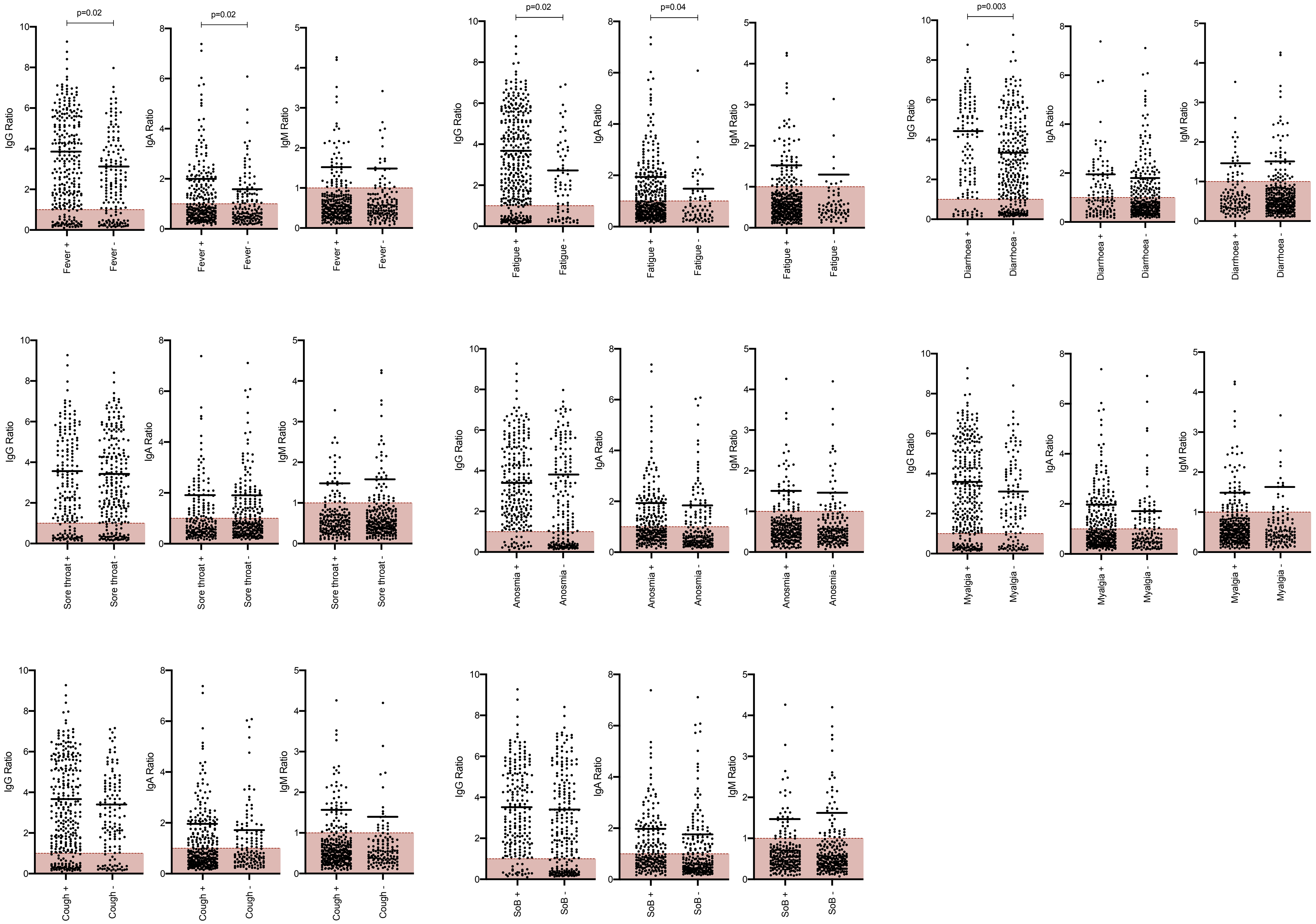


Supplementary
Figure 3

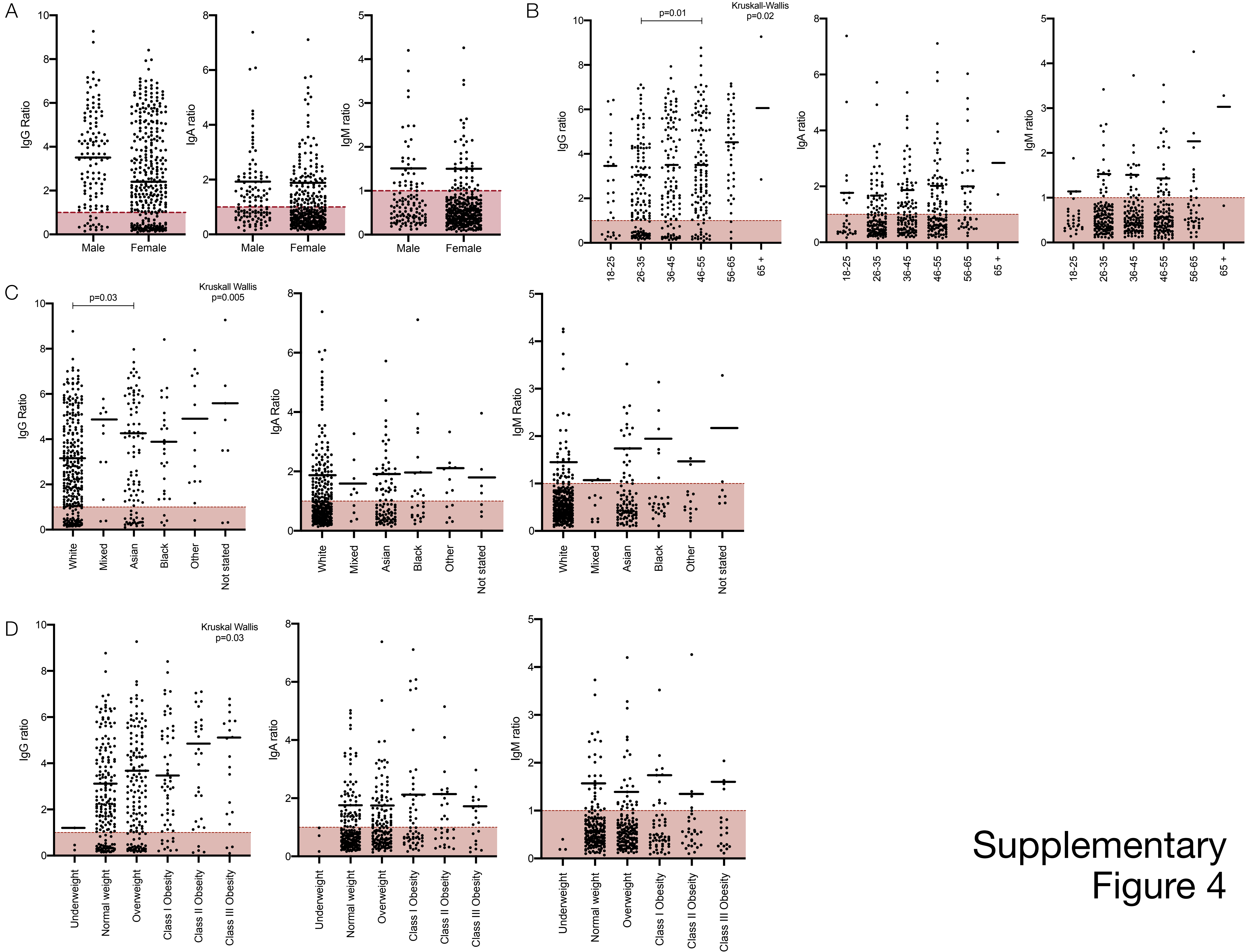


Supplementary Figure 4
