## Supplementary Tables for "Serological responses to SARS-CoV-2 following non-hospitalised infection: clinical and ethnodemographic features associated with the magnitude of the antibody response"

| **Variable** | **OR (95% CI)** | **Z** | **P-value** |
| --- | --- | --- | --- |
| Age (years) | 0.99  (0.97-1.01) | 0.91 | 0.37 |
| Sex (Female) | 0.78 (0.42-1.41) | 0.83 | 0.41 |
| **Ethnicity (BAME)** | **1.93 (1.11-3.40)** | **2.31** | **0.02** |
| Job role : Administration | 0.42  (0.07-2.20) | 1.00 | 0.32 |
| Job role : AHP | 0.41 (0.07-1.92) | 1.09 | 0.27 |
| Job role : Doctor | 0.46 (0.09-1.91) | 1.02 | 0.31 |
| Job role: HCA | 0.61 (0.10-3.11) | 0.58 | 0.56 |
| Job role : Lab scientist | 1.10 (0.15-7.96) | 0.10 | 0.92 |
| Job role: Managerial | 0.19 (0.02-1.22) | 1.68 | 0.09 |
| Job role : Nurse/Midwife | 0.61  (0.12-2.43) | 0.67 | 0.50 |
| Job role : Research nurse | 0.26 (0.04-1.44) | 1.49 | 0.14 |

**Supplementary Table 1:**  Multiple logistic regression of seropositivity at time of study enrolment with respect to job role. Area under the curve for this model was 0.61 (CI 0.55-0.68, p=0.0008).

| **Variable** | **OR (95% CI)** | **Z** | **P-value** |
| --- | --- | --- | --- |
| Age (years) | 0.99  (0.97-1.01) | 0.57 | 0.57 |
| Sex (Female) | 0.8  (0.49-1.40) | 0.70 | 0.48 |
| **Ethnicity (BAME)** | **1.93**  **(1.10-3.43)** | **2.27** | **0.02** |
| Department: Admin/management | 0.54  (0.14-1.97) | 0.92 | 0.36 |
| Department:  AMU | 1.09  (0.32-3.85) | 0.14 | 0.89 |
| Department:  ED | 2.05  (0.53-9.05) | 1.01 | 0.31 |
| Department: Endoscopy/Theatres | 1.07  (0.26-4.57) | 0.09 | 0.93 |
| Department: GIM/Geriatrics | 1.43  (0.59-3.45) | 0.80 | 0.42 |
| Department: Haematology/Oncology | 2.67  (0.64-14.05) | 1.28 | 0.20 |
| Department: ITU | 1.23  (0.45-3.38) | 0.40 | 0.69 |
| Department: Laboratory medicine | 1.35  (0.33-6.16) | 0.41 | 0.68 |
| Department: Pharmacy | 0.51  (0.12-1.94) | 0.97 | 0.33 |
| Department: R&D | 0.3  (0.04-1.59) | 1.32 | 0.19 |
| Department: Renal | 0.67  (0.20-2.16) | 0.67 | 0.50 |
| Department: Surgery | 0.98  (0.36-2.67) | 0.03 | 0.97 |

**Supplementary Table 2:**  Multiple logistic regression of seropositivity at time of study enrolment with respect to department. Area under the curve for this model was 0.64 (CI 0.58-0.71, p<0.0001).
